# Seasonally Normalized Vitamin D Status and Long-Term Mortality and Multisystem Disease Risk in the UK Biobank

**DOI:** 10.64898/2026.10.07.26364983

**Authors:** Ariel Israel, Abraham Weizman, Shai Ashkenazi, Eli Magen, Eugene Merzon, Shlomo Vinker, Nadav Rappoport

## Abstract

Vitamin D deficiency has been associated with mortality and chronic disease. In a recent study of Israeli Leumit Health Services (LHS) and US TriNetX populations, severe deficiency was reproducibly associated with mortality and multisystem morbidity, while longitudinal supplementation reconstructed from pharmacy dispensing was associated with lower risk.

**Methods:** We sought to reproduce these associations and address a wide range of potential confounders in the deeply phenotyped UK Biobank. Baseline circulating 25-hydroxyvitamin D [25(OH)D] was standardized to a common late-winter nadir using an eight-harmonic model, and participants were categorized according to nadir-equivalent 25(OH)D as <5, 5-10, 10-15, 15-20, or ≥20 ng/mL. Primary survival analyses used one-year delayed entry and complete-case multivariable adjustment.

**Results:** Analyses included 412,020 participants and 31,197 deaths. Mortality hazard ratios were 2.03 (95% CI 1.90-2.18), 1.34 (1.29-1.39), 1.08 (1.04-1.11), respectively, for <5, 5-10 and 10-15, versus ≥20 ng/mL. Associations previously observed in LHS and TriNetX were reproduced, notably for diabetes, myocardial infarction, cerebrovascular disease, dementia, diabetic retinopathy, and end-stage renal disease, whereas skin malignancies showed the opposite gradient, consistent with greater ultraviolet exposure. DAG-informed causal analyses using generalized overlap weighting achieved acceptable balance across measured covariates; weighted 10-year mortality was 9.7% with very severe deficiency versus 4.8% with ≥20 ng/mL, supporting a causal interpretation. Estimates remained stable with landmark lags extending to 5 years after measurement.

**Conclusions:** Seasonally normalized 25(OH)D identifies a reproducible state of systemic vulnerability while reducing exposure misclassification caused by calendar timing of measurement. An interactive implementation of the normalization model is available: https://vitamind.medil.ai.

## 1. Introduction

Vitamin D deficiency is common worldwide and varies substantially with latitude, skin pigmentation, lifestyle, adiposity, and ultraviolet exposure (Cashman et al. 2016; Hilger et al. 2014; Holick 2007). Beyond its established role in skeletal health, low circulating 25-hydroxyvitamin D [25(OH)D] has repeatedly been associated with mortality and disorders spanning metabolic, cardiovascular, renal, microvascular, and neurological domains (Afzal et al. 2013; Gotsman et al. 2012; Judd & Tangpricha 2009; Usluogullari et al. 2015). Whether these associations reflect a biological contribution of vitamin D, reverse causation from underlying disease, or confounding by the many determinants of vitamin D status remains debated (Manson et al. 2019; Neale et al. 2022).

In a previous study of two large adult healthcare populations, Leumit Health Services (LHS) in Israel and the US TriNetX network, severe vitamin D deficiency was reproducibly associated with increased mortality and multisystem morbidity (Israel, Weizman, et al. 2026). Importantly, vitamin D supplementation reconstructed longitudinally from pharmacy-dispensed purchases and calibrated against subsequent serum measurements remained independently and dose-dependently associated with lower mortality and morbidity, including in causal-framework analyses accounting for time-varying treatment selection. These findings supported vitamin D deficiency as a potentially modifiable risk state.

However, because these analyses were based on electronic health records, information was limited for several important phenotypic and environmental determinants of vitamin D status, including ethnicity, skin colour, socioeconomic deprivation, and outdoor exposure. Imbalances in these factors could contribute to residual confounding in associations based on isolated 25(OH)D measurements. A further important limitation is seasonal exposure misclassification. At temperate latitudes, circulating 25(OH)D follows a pronounced annual cycle (Hyppönen & Power 2007). Consequently, the same measured concentration can represent substantially different underlying vitamin D states depending on when during the year it is obtained. Fixed thresholds applied without reference to test date can therefore combine individuals with different annual vitamin D trajectories within the same exposure category, while separating individuals with similar trajectories into different categories.

The deeply phenotyped UK Biobank provides an opportunity to address these limitations more comprehensively. Its systematic characterization of demographic, behavioural, socioeconomic, geographic, pigmentation-related, environmental, and clinical factors, together with documented timing of blood sampling and longitudinal outcome ascertainment, allows substantially broader control of potential confounding and direct modelling of seasonal variation (Bycroft et al. 2018; Sudlow et al. 2015).

We therefore modelled the annual 25(OH)D cycle and transformed each baseline measurement to a common late-winter nadir-equivalent concentration before assigning vitamin D categories. We then examined mortality and multisystem disease outcomes, compared the information provided by raw and seasonally normalized measurements, and specified a directed acyclic graph (DAG) to identify major measured common causes of vitamin D status and mortality. Generalized overlap weighting was used to test whether the mortality association persisted after achieving covariate balance across vitamin D strata (Greenland et al. 1999; Li et al. 2018; Li & Li 2019).

We hypothesized that severe nadir-equivalent vitamin D deficiency would reproduce the mortality and multisystem associations observed previously and that the severe-deficiency mortality association would persist after balancing the major measured determinants of both vitamin D status and mortality. The causal and measurement framework is summarized in Figure 1.

**Figure 1.**
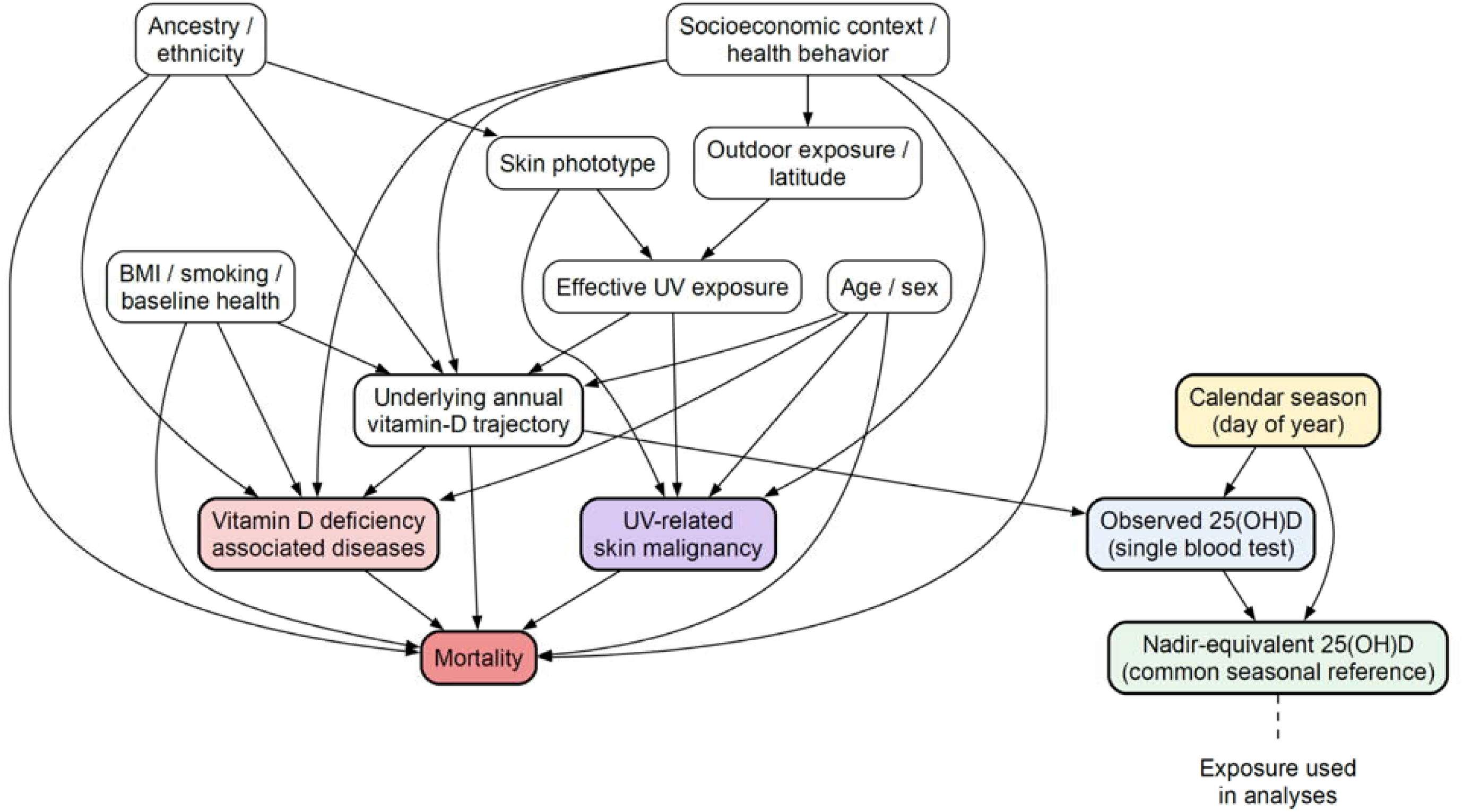
Causal and measurement framework for vitamin-D status, disease outcomes, skin malignancy, and mortality. Directed acyclic graph representing the hypothesized relationships among demographic, ancestry/ethnicity, socioeconomic, behavioral, metabolic, photobiological, geographic, and baseline-health determinants of the underlying annual vitamin-D trajectory and downstream outcomes. Ultraviolet exposure contributes both to vitamin-D status and to UV-related skin malignancy. Vitamin D deficiency-associated disease outcomes and skin malignancy may in turn contribute to mortality. The measurement pathway distinguishes the underlying annual vitamin-D trajectory from the observed single 25(OH)D blood test, calendar season, and the derived nadir-equivalent 25(OH)D value used as the standardized exposure in the analyses. Directed edges encode the prespecified causal assumptions used to identify the measured adjustment set and to distinguish the underlying exposure from its measurement process.

## 2. Materials and Methods

### 2.1 UK Biobank cohort

UK Biobank is a large prospective cohort with extensive baseline phenotyping and longitudinal linkage to health outcomes (Bycroft et al., 2018; Sudlow et al., 2015). We included participants with an available baseline serum 25(OH)D measurement and the clinical data required for each analysis. The vitamin D source population comprised 447,842 participants. UK Biobank data were accessed under Application No. 56774 and extracted in October 2025. Further details of cohort construction, variable derivation, data cleaning, and analytical implementation are provided in the Supplementary Methods.

### 2.2 Exposure definition and seasonal normalization

The observed exposure was baseline serum 25(OH)D concentration (UK Biobank field 30890), converted from nmol/L to ng/mL by division by 2.5. Calendar day of measurement was represented using eight sine/cosine harmonic pairs, allowing a flexible smooth periodic annual pattern rather than imposing a single sinusoid (Stolwijk et al. 1999). The seasonal component was defined as:

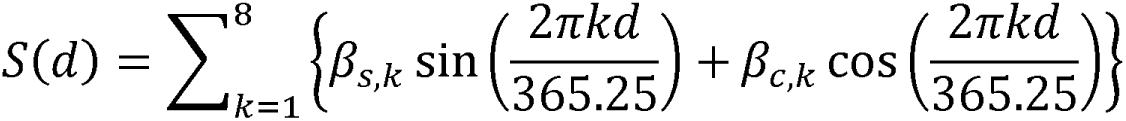

Coefficients were estimated by linear regression of log-transformed baseline 25(OH)D on the 16 harmonic terms in the full vitamin D cohort. For participant *i* measured on day d_i_, S(d*_i_*), represents the fitted seasonal component on the measurement day. The fitted annual nadir occurred on day 59 (late February), with *S*(*d_nadir_*) representing the corresponding fitted seasonal component. Nadir-equivalent 25(OH)D was then defined as:

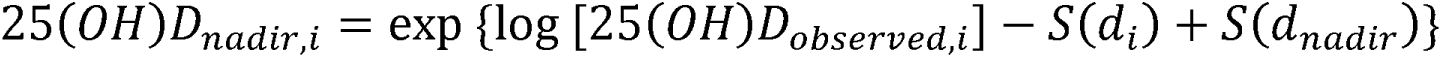

This transformation removes the fitted calendar component from the observed measurement and expresses each value at a common late-winter reference.

Nadir-equivalent and raw 25(OH)D measurements were categorized using the same thresholds: <5, 5–10, 10–15, 15–20, and ≥20 ng/mL. The ≥20 ng/mL nadir-equivalent category served as the primary reference. Raw categories were defined by applying the same numerical thresholds directly to the observed laboratory measurement.

The complete harmonic coefficients used for the submitted analyses are provided in the Supporting Information together with the transformation specification, allowing the nadir-equivalent exposure to be reproduced independently of the interactive web implementation.

An interactive implementation of the seasonal normalization model, allowing conversion of an observed 25(OH)D concentration and measurement date to its nadir-equivalent value, is available at https://vitamind.medil.ai. The interactive calculator provides a user-facing implementation of the same normalization model.

### 2.3 Baseline covariates and determinant analysis

The primary association models adjusted for age, sex, broad self-reported ethnicity, BMI category and within-category BMI offset, baseline diabetes, current and past smoking, skin colour, reported summer and winter outdoor exposure, Townsend deprivation index, geographic position, calendar year of baseline assessment, and fish-oil use. Source fields and detailed recoding rules for all baseline covariates are provided in the Supplementary Methods. Broad ethnicity was derived from self-reported ethnic background and categorized as White, Asian, Black, Mixed, Chinese, or Other; responses indicating “do not know,” “prefer not to answer,” or unknown ethnicity were treated as missing. BMI was categorized as 10–18.5, 18.5–25, 25–30, 30–35, 35–40, 40–45, or 45–99 kg/m², with 18.5–25 kg/m² as the reference category; a within-category BMI offset retained continuous BMI information within each category. Baseline diabetes was defined by an E10 or E11 first-report date on or before baseline. Sex was omitted from sex-specific models.

Fish-oil use was derived from baseline supplement field 6179 (code 1). Vitamin D-containing medications were additionally sought in baseline medication field 20003 but showed no informative variation in the available dataset and therefore did not enter the fitted models.

UK Biobank special response codes were cleaned before analysis. For example, responses of ‘do not know’ and ‘prefer not to answer’ for outdoor time were coded as missing, whereas ‘less than an hour a day’ was coded as 0.5 h/day.

### 2.4 Outcomes and follow-up

The primary outcome was all-cause mortality. Main secondary outcomes, analyzed separately, were incident diabetes mellitus, diabetic retinopathy, myocardial infarction, cerebrovascular infarction, dementia, and end-stage renal disease, selected because they directly replicated disease domains identified in the preceding LHS/TriNetX study (Israel, Weizman, et al. 2026). Melanoma and other skin malignancies were analyzed as ultraviolet-related photobiological outcomes. Operational definitions of all study outcomes, including the corresponding UK Biobank source fields, ICD-10 codes, cancer-registry definitions, and construction of composite outcomes, are provided in Supplementary Table S1.

Time zero was the baseline UK Biobank assessment, at which 25(OH)D was measured. For each outcome, follow-up ended at the earliest of outcome occurrence, death for nonfatal outcomes, loss to follow-up, or the global data-censoring date, defined as one day after the latest recorded death date in the dataset. Follow-up was additionally capped at 13.1 years from each participant’s baseline assessment, corresponding to the maximum analytically usable follow-up duration. Primary analyses used delayed entry at one year after baseline to reduce potential reverse causation; participants experiencing the outcome or being censored before this point did not contribute to the analysis, while baseline remained the time origin.

### 2.5 Conventional Cox models and standardized curves

Cox proportional-hazards models were fitted with vitamin D category and the prespecified covariates using the follow-up and censoring definitions described above. Sex-specific models omitted sex from the adjustment set. Adjusted hazard ratios (HRs) and 95% confidence intervals (CIs) were derived from the fitted models. Model-based cumulative-incidence curves were obtained using a common covariate profile, thereby displaying differences between vitamin D categories at matched values of the measured covariates. For nonfatal outcomes, these curves represent Cox model-based cumulative event probabilities rather than competing-risk cumulative-incidence functions.

Proportional-hazards assumptions were evaluated using scaled Schoenfeld residuals. In sparse outcome/subgroup combinations where full cox.zph diagnostics were numerically singular, exposure-focused time-interaction diagnostics were used as a targeted sensitivity analysis rather than treating a diagnostic failure as an outcome-model failure.

### 2.6 Raw-versus-nadir model comparison

For every outcome and sex stratum, covariate-only, raw-vitamin-D, and nadir-equivalent-vitamin-D Cox models were fitted on an identical complete-case population. Model fit was compared using the Akaike information criterion (AIC) and concordance. Time-dependent area under the receiver operating characteristic curve (AUC), accounting for right censoring, was estimated at 2, 3, 4, 5, and 10 years from baseline measurement. We also quantified overall and month-specific movement between raw and nadir-equivalent categories.

### 2.7 DAG-informed causal mortality analysis

A directed acyclic graph (Figure 1) was specified to formalize the hypothesized common causes of vitamin-D status, disease outcomes, and mortality and to distinguish the underlying annual vitamin-D trajectory from the season-dependent laboratory measurement (Greenland et al. 1999). The measured causal adjustment set comprised age, sex, broad ethnicity, BMI category and within-category BMI, baseline diabetes, current and past smoking, skin colour, summer and winter outdoor exposure, Townsend deprivation, geographic position, calendar year, fish-oil use, and baseline histories of myocardial infarction, congestive heart failure, cerebral infarction, chronic kidney disease/end-stage renal disease, cirrhosis, dementia, and cancer. Vitamin D deficiency-associated incident diseases and UV-related skin malignancy were depicted as downstream outcomes that can contribute to mortality; they were not entered as time-varying mortality covariates because doing so could condition on mediators of the total vitamin-D effect.

### 2.8 Generalized overlap weighting and absolute risks

Generalized propensity scores for the five nadir-equivalent categories were estimated using multinomial regression. Generalized overlap weights were calculated from these scores to target the population with the greatest covariate overlap across all exposure categories (Li et al. 2018; Li & Li 2019). Balance was summarized by the maximum absolute pairwise SMD for each covariate before and after weighting, with <0.10 prespecified as acceptable.

Weighted Cox models used robust variance estimation clustered by participant identifier. Weighted survival functions were used to estimate mortality risks at 5 and 10 years from baseline, together with corresponding risk differences and risk ratios. Reverse-causation sensitivity analyses repeated the complete weighting procedure using landmark lags of 0, 1, 2, 3, 4, and 5 years.

E-values were calculated for the primary severe-deficiency effect estimate and confidence-limit closest to the null (VanderWeele & Ding 2017). Causal interpretation was based on the standard assumptions of consistency, conditional exchangeability given the measured covariates, positivity, and correct model specification (Cole & Hernán 2008).

### 2.9 Statistical analysis

Continuous variables are summarized as mean ± SD where appropriate and categorical variables as counts and percentages. Table 1 emphasizes standardized mean differences rather than significance testing because of the large cohort size. For secondary outcomes, Benjamini– Hochberg false-discovery-rate adjustment was applied to the prespecified vitamin-D contrasts. All analyses were performed using R (version 4) and Python (3.13).

**Table 1.** Baseline characteristics according to nadir-equivalent 25(OH)D category.

| Variable | Overall | 0–5<br>ng/mL | 5–10<br>ng/mL | 10–15<br>ng/mL | 15–20<br>ng/mL | ≥20<br>ng/mL | SMD |
| --- | --- | --- | --- | --- | --- | --- | --- |
| N | 447,842 | 9,363 | 92,551 | 144,365 | 115,359 | 86,204 | — |
| Age, years | 56.48<br>(8.12) | 53.80<br>(8.17) | 55.20<br>(8.14) | 56.30<br>(8.12) | 57.03<br>(8.07) | 57.69<br>(7.89) | 0.238 |
| Female, n (%) | 239,818<br>(53.5) | 4,989<br>(53.3) | 50,617<br>(54.7) | 76,450<br>(53.0) | 60,409<br>(52.4) | 47,353<br>(54.9) | 0.028 |
| White ethnicity, n (%) | 422,691<br>(94.8) | 6,049<br>(65.6) | 81,507<br>(88.7) | 137,875<br>(95.9) | 112,518<br>(97.9) | 84,742<br>(98.6) | 0.476 |
| BMI, kg/m<sup>2</sup> | 27.41<br>(4.79) | 28.86<br>(6.27) | 28.50<br>(5.58) | 27.69<br>(4.77) | 26.92<br>(4.28) | 26.28<br>(3.99) | 0.270 |
| Diabetes, n (%) | 13,836<br>(3.1) | 651<br>(7.0) | 4,142<br>(4.5) | 4,393<br>(3.0) | 2,768<br>(2.4) | 1,882<br>(2.2) | 0.116 |
| Current smoker, n (%) | 34,399<br>(7.7) | 1,716<br>(18.3) | 11,037<br>(11.9) | 10,389<br>(7.2) | 6,479<br>(5.6) | 4,778<br>(5.5) | 0.210 |
| Fair skin, n (%) | 300,529<br>(67.1) | 4,628<br>(49.4) | 58,711<br>(63.4) | 98,841<br>(68.5) | 79,495<br>(68.9) | 58,854<br>(68.3) | 0.405 |
| Summer outdoor time, h/day | 3.80<br>(2.40) | 3.05<br>(2.35) | 3.31<br>(2.33) | 3.67<br>(2.36) | 4.00<br>(2.39) | 4.31<br>(2.44) | 0.270 |
| Winter outdoor time, h/day | 1.95<br>(1.82) | 1.77<br>(1.89) | 1.77<br>(1.77) | 1.91<br>(1.79) | 2.05<br>(1.84) | 2.09<br>(1.85) | 0.102 |
| Townsend deprivation index | −1.31<br>(3.08) | 0.66<br>(3.56) | −0.54<br>(3.38) | −1.36<br>(3.03) | −1.69<br>(2.87) | −1.78<br>(2.81) | 0.376 |
| Fish-oil use, n (%) | 139,920<br>(31.2) | 983<br>(10.5) | 15,058<br>(16.3) | 39,238<br>(27.2) | 44,417<br>(38.5) | 40,224<br>(46.7) | 0.446 |

Values are mean (SD) unless otherwise indicated. SMD is the overall standardized mean difference across the five vitamin-D categories and summarizes baseline differences across the five vitamin D categories.

Additional methodological details, including seasonal-model coefficients, covariate derivation, complete-case preparation, censoring procedures, proportional-hazards diagnostics, weighting diagnostics, and causal sensitivity analyses, are provided in the Supplementary Methods.

### 2.10 Use of generative artificial intelligence

During manuscript preparation, the authors used ChatGPT (OpenAI) for assistance with language editing and manuscript organization. The tool was not used to generate the underlying study data or perform the statistical analyses. All AI-assisted content was critically reviewed, verified, and edited by the authors, who take full responsibility for the final manuscript.

### 2.11 Ethics and data availability

UK Biobank operates under approval from the North West Multi-centre Research Ethics Committee as a Research Tissue Bank (REC reference 11/NW/0382), and all participants provided informed consent. The present study was conducted under UK Biobank Application Number 56774. The participant-level data used in this study are available to approved researchers through UK Biobank and cannot be redistributed by the authors.

## 3. Results

### 3.1 Study population and determinants of vitamin D status

The source population comprised 447,842 participants with a baseline 25(OH)D measurement. Mean age was 56.48 years (SD 8.12), 53.5% were female, and 94.8% were classified within the broad White ethnicity category. Nadir-equivalent 25(OH)D categorized 9,363 participants as 0–5 ng/mL, 92,551 as 5–10 ng/mL, 144,365 as 10–15 ng/mL, 115,359 as 15–20 ng/mL, and 86,204 as ≥20 ng/mL (Table 1).

Vitamin D categories differed markedly in characteristics expected to influence cutaneous synthesis, behavior, and general health. Participants in the 0–5 ng/mL group were younger on average than those with ≥20 ng/mL (53.80 vs. 57.69 years), had higher BMI (28.86 vs. 26.28 kg/m²), were more frequently current smokers, reported less summer outdoor exposure (3.05 vs. 4.31 h/day), and had greater socioeconomic deprivation. Ethnic and pigmentation differences were particularly pronounced: 65.6% of the 0–5 ng/mL group versus 98.6% of the ≥20 ng/mL group were categorized as White, and 49.4% versus 68.3% had fair skin. These differences illustrate the extensive host, behavioral, and environmental structure underlying an apparently simple laboratory measurement.

### 3.2 Calendar normalization places measurements on a common seasonal scale

Observed 25(OH)D showed a strong annual cycle, with lowest concentrations in late winter and highest concentrations in late summer. We modeled this pattern on the log scale using eight sine/cosine harmonic pairs and transformed each participant’s observed baseline measurement to the estimated concentration corresponding to the modeled annual nadir. The resulting variable is a calendar-season-normalized, nadir-equivalent estimate. Figure 2 summarizes the seasonal model and nadir nomogram (A–B), month-specific category reclassification (C), and selected independent determinants of 25(OH)D (D).

**Figure 2.**
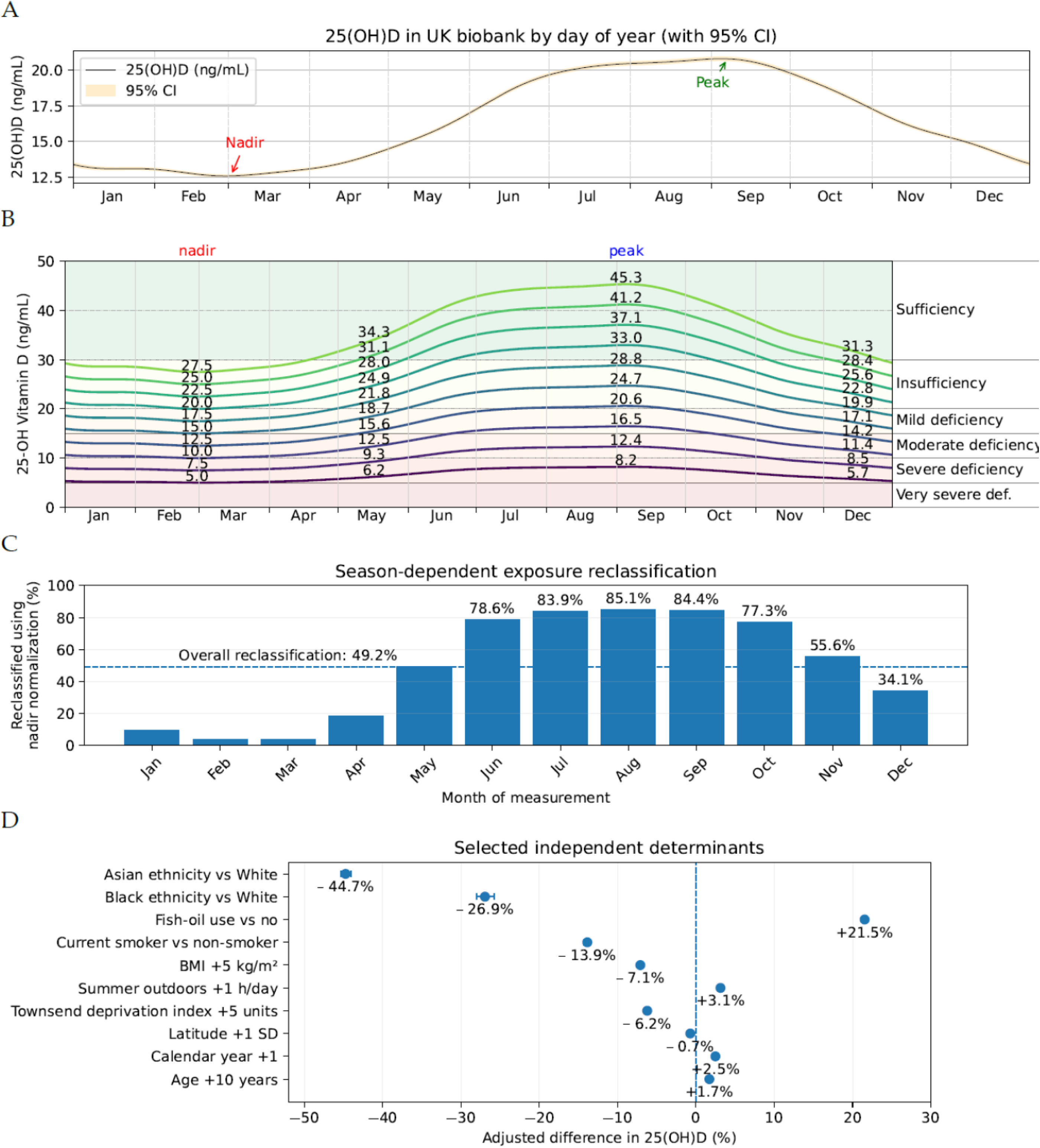
Seasonal variation, nadir normalization, exposure reclassification, and independent determinants of 25(OH)D. (A) Observed annual 25(OH)D pattern and fitted eight-harmonic seasonal function, showing the late-winter nadir and late-summer peak. (B) Seasonal nomogram showing annual trajectories corresponding to specified nadir-equivalent 25(OH)D concentrations and illustrating that approximately 20 ng/mL at the nadir corresponds to approximately 33 ng/mL near the seasonal peak. (C) Percentage of participants whose vitamin-D category changed after normalization to the common seasonal nadir, by month of measurement; overall reclassification was 49.2% and exceeded 80% around the late-summer peak. (D) Selected independent determinants of log-transformed 25(OH)D from the multivariable determinant model, expressed as adjusted percentage differences for the contrasts/scales shown. The nadir-equivalent value is modeled, not directly observed.

**Figure 3.**
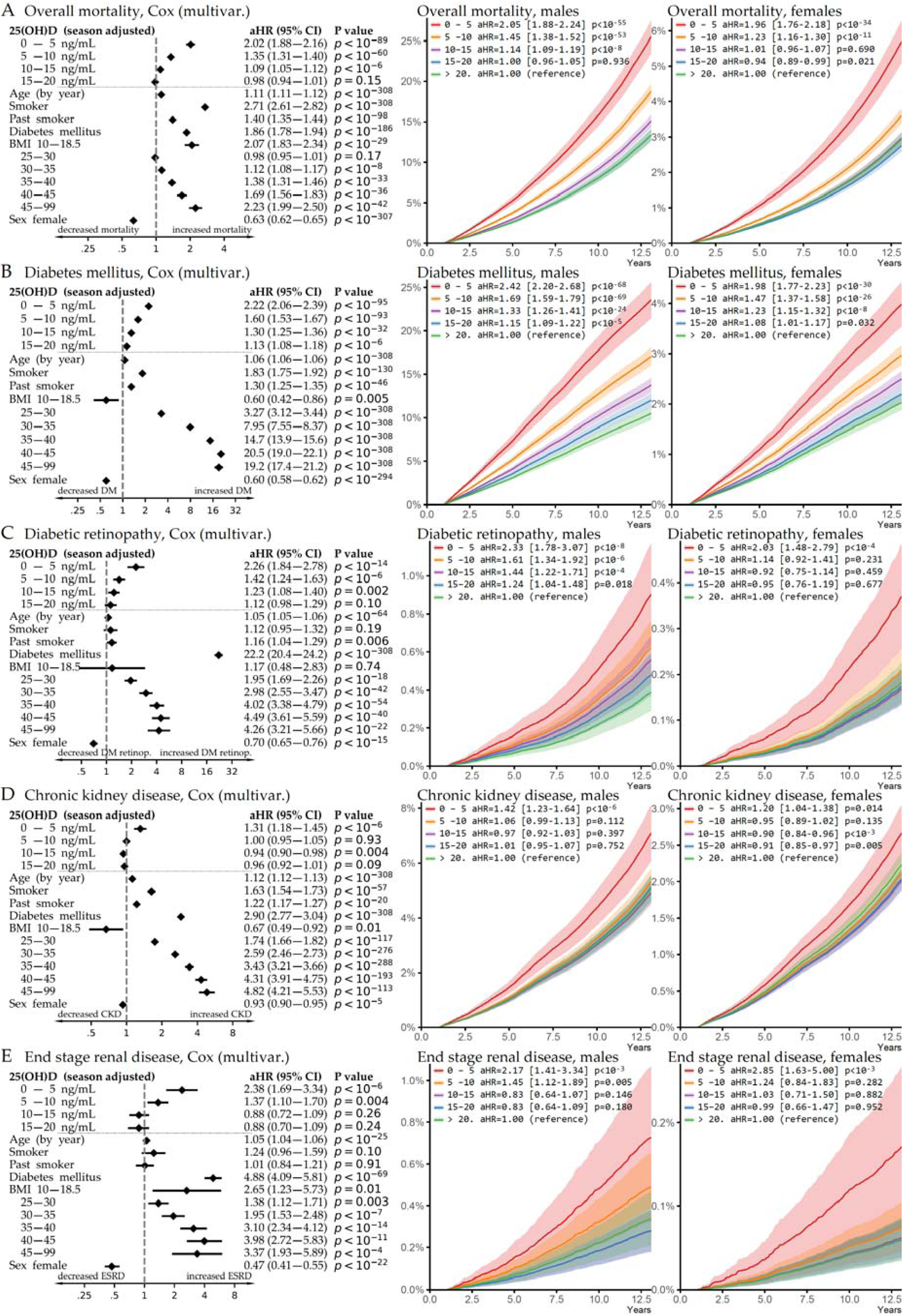
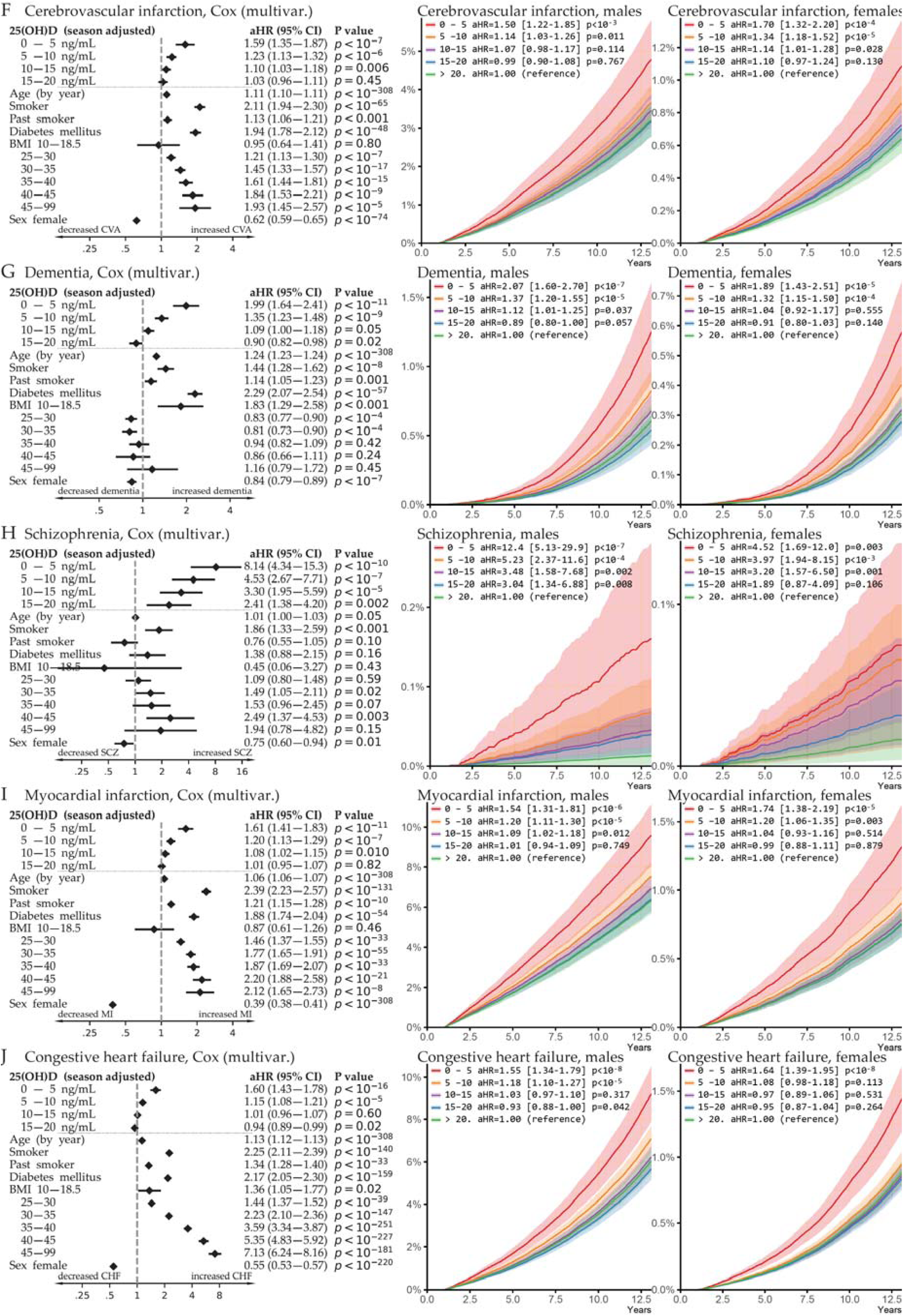
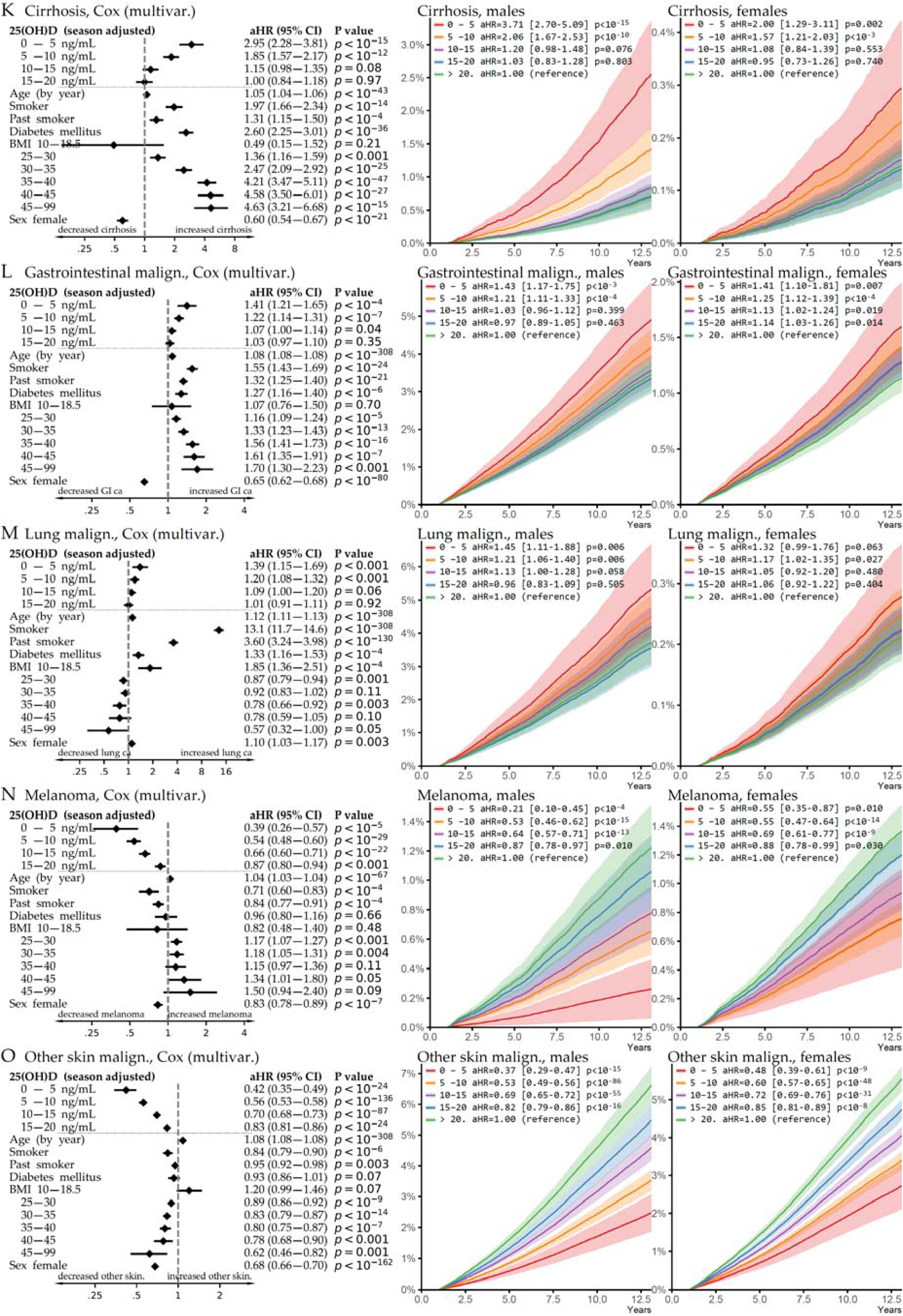
Multivariable associations of nadir-equivalent 25(OH)D with mortality and multisystem disease outcomes. Multivariable associations of nadir-equivalent 25(OH)D with mortality and multisystem disease outcomes. For each outcome, the left plot displays the complete vitamin D dose-response together with selected covariate estimates from the fully adjusted model; all models additionally included the prespecified demographic, pigmentation-related, environmental, socioeconomic, and geographic covariates described in Methods. The center and right plots show sex-specific standardized cumulative-incidence curves derived from the fitted Cox models. These curves are model-based projections rather than unadjusted Kaplan–Meier curves. The ≥20 ng/mL nadir-equivalent category is the vitamin D reference.

**Figure 4.**
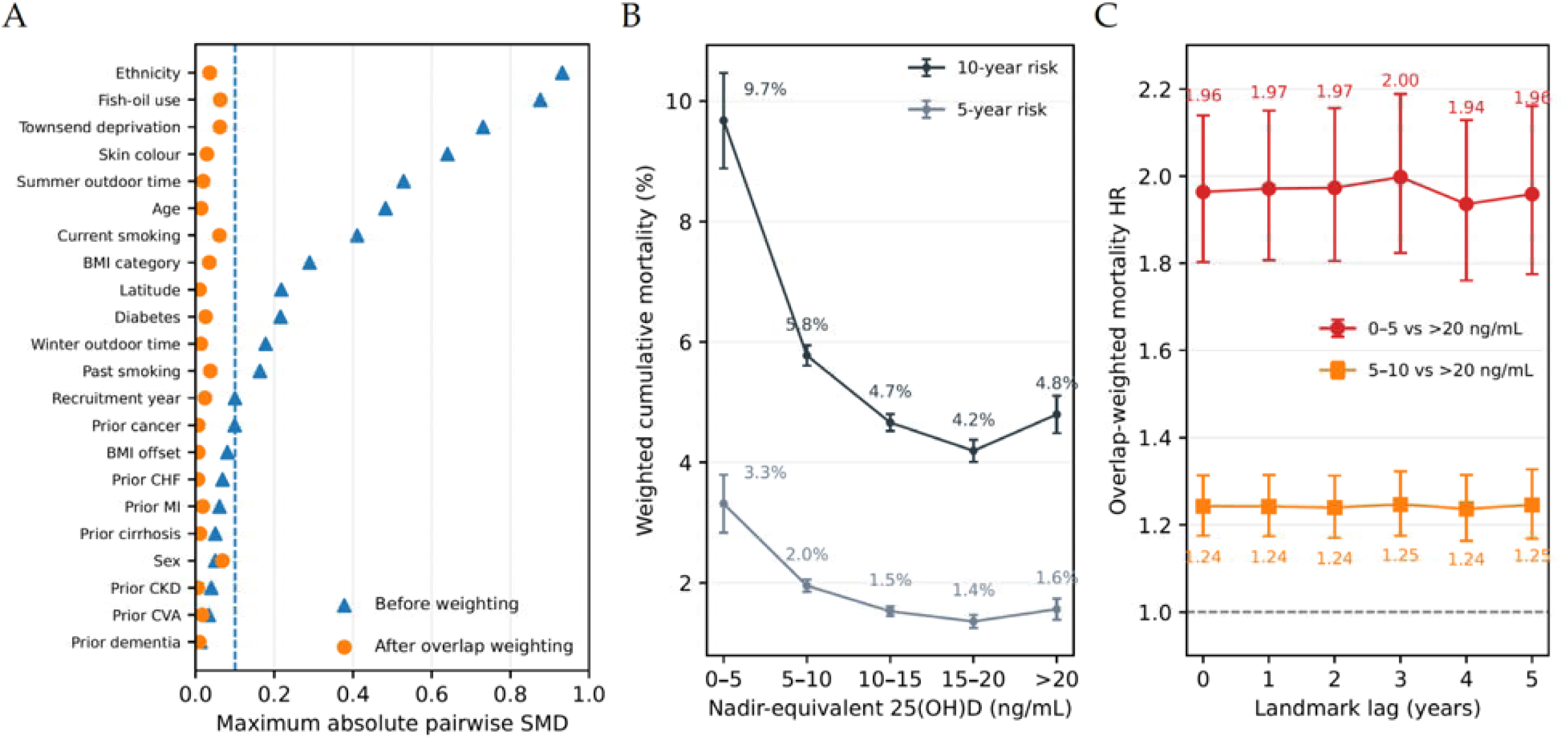
DAG-informed causal analysis of nadir-equivalent 25(OH)D and all-cause mortality. (A) Maximum absolute pairwise standardized mean differences across the five nadir-equivalent 25(OH)D categories before and after generalized overlap weighting at the primary one-year delayed-entry landmark. (B) Overlap-weighted absolute cumulative mortality at 5 and 10 years from baseline across the five vitamin D categories; 10-year mortality was 9.7% for <5 ng/mL versus 4.8% for ≥20 ng/mL. (C) Sensitivity of the overlap-weighted mortality associations for <5 and 5–10 ng/mL, each versus ≥20 ng/mL, to delayed-entry lags from 0 through 5 years. Corresponding HRs remained approximately 1.94–2.00 and 1.24–1.25, respectively, with no material attenuation as the lag increased.

Seasonal normalization changed the vitamin D category of 220,300 of 447,842 participants (49.2%). Reclassification was minimal close to the nadir, affecting approximately 3.7% of February and March measurements, but increased sharply during the seasonal peak: 78.6% in June, 83.9% in July, 85.1% in August, and 84.4% in September. This strong calendar dependence is the pattern expected when fixed thresholds are applied to a biomarker with substantial predictable annual displacement.

### 3.3 Nadir-equivalent vitamin D and all-cause mortality

After one-year delayed entry, the primary mortality analysis included 412,020 participants and 31,197 deaths. Relative to nadir-equivalent 25(OH)D ≥20 ng/mL, adjusted hazard ratios were 2.03 (95% CI 1.90–2.18) for 0–5 ng/mL, 1.34 (1.29–1.39) for 5–10 ng/mL, 1.08 (1.04–1.11) for 10–15 ng/mL. The steepest increase therefore occurred below 10 ng/mL, and particularly below 5 ng/mL, with little evidence of additional mortality reduction above approximately 15–20 ng/mL when expressed at the annual nadir.

The pattern was present in both sexes but was quantitatively stronger in men (global vitamin D-by-sex interaction P=8.0×10). In men, HRs for 0–5 and 5–10 ng/mL were 2.08 (1.90–2.28) and 1.43 (1.37–1.50); corresponding estimates in women were 1.98 (1.78–2.21) and 1.22 (1.15–1.30).

### 3.4 Replication across major disease domains

The multisystem pattern previously identified in LHS and TriNetX was independently reproduced in UK Biobank. Incident diabetes showed a graded association across the full low-vitamin-D range, with adjusted HRs of 1.80 (1.66–1.95), 1.50 (1.43–1.57), 1.27 (1.22–1.33), and 1.12 (1.07–1.17) across the four sub-reference categories. Diabetic retinopathy was similarly increased, with HRs of 2.00 (1.62–2.49), 1.37 (1.19–1.57), 1.22 (1.07–1.39), and 1.13 (0.98–1.29).

Cardiovascular and cerebrovascular outcomes reproduced the same direction. Myocardial infarction HRs were 1.49 (1.30–1.71), 1.18 (1.10–1.26), 1.07 (1.01–1.14), and 1.00 (0.94–1.06), while cerebrovascular infarction HRs were 1.58 (1.34–1.86), 1.23 (1.13–1.33), 1.11 (1.03–1.19), and 1.03 (0.96–1.11). Dementia showed a particularly strong association at very low concentrations: HR 2.08 (1.71–2.53) for 0–5 ng/mL and 1.37 (1.25–1.51) for 5–10 ng/mL. End-stage renal disease was likewise increased at the lowest concentrations, with HR 2.12 (1.49–3.02) for 0–5 ng/mL and 1.29 (1.03–1.61) for 5–10 ng/mL.

### 3.5 Ultraviolet-related skin malignancy shows the opposite gradient

As in the preceding LHS and TriNetX analyses, melanoma and other skin malignancies were examined as photobiological control outcomes because ultraviolet exposure increases both cutaneous vitamin D synthesis and skin-cancer risk. The UK Biobank results reproduced the expected inverse gradient. After adjustment for the core and ultraviolet-related covariates, melanoma HRs were 0.43 (95% CI 0.29–0.63), 0.54 (0.48–0.60), 0.65 (0.60–0.71), and 0.87 (0.80–0.94) across the <5, 5–10, 10–15, and 15–20 ng/mL categories, respectively, relative to ≥20 ng/mL. Corresponding HRs for other skin malignancies were 0.46 (0.39–0.55), 0.56 (0.54– 0.59), 0.70 (0.67–0.72), and 0.83 (0.80–0.86). Thus, skin malignancies behaved in the opposite direction from mortality and systemic disease, consistent with the shared influence of ultraviolet radiation on vitamin D synthesis and photodamage (Armstrong & Kricker 2001; Gandini et al. 2005).

### 3.6 DAG-informed overlap weighting removes the major measured differences between vitamin D strata

We next tested whether the mortality association persisted after achieving covariate comparability according to the prespecified causal framework (Figure 1). Generalized overlap weighting was selected because the five exposure categories showed areas of limited common support and conventional inverse-probability weighting can become unstable in such settings (Li et al. 2018; Li & Li 2019). Before weighting, maximum pairwise SMDs reached 0.93 for ethnicity, 0.87 for fish-oil use, 0.73 for deprivation, 0.64 for skin colour, 0.53 for summer outdoor exposure, and 0.49 for age. At the one-year landmark, overlap weighting reduced every measured covariate to a maximum pairwise SMD <0.10; the largest residual SMD was approximately 0.068, with ethnicity 0.035, skin colour 0.028, summer outdoor exposure 0.019, and age 0.013.

### 3.7 Severe deficiency remains associated with mortality after overlap weighting

Even after achieving measured-covariate balance, the mortality association persisted. At the primary one-year landmark, overlap-weighted HRs were 1.97 (1.81–2.15) for 0–5 ng/mL and 1.24 (1.17–1.31) for 5–10 ng/mL.

Absolute risks were similarly informative. At 10 years from baseline, weighted mortality was 9.7% in the <5 ng/mL category, 5.8% at 5–10 ng/mL, 4.7% at 10–15 ng/mL, 4.2% at 15–20 ng/mL, and 4.8% in the ≥20 ng/mL reference. Corresponding 5-year risks were 3.3%, 2.0%, 1.5%, 1.4%, and 1.6%. The mortality associations were remarkably stable under progressively longer landmark exclusions. Across 0-, 1-, 2-, 3-, 4-, and 5-year landmarks, the overlap-weighted HR for 0–5 versus ≥20 ng/mL remained between 1.94 and 2.00, while the HR for 5–10 versus ≥20 ng/mL remained between 1.24 and 1.25. This absence of material attenuation argues against early preclinical illness as the primary explanation for the observed excess risk.

The E-value for the primary 0–5 ng/mL estimate was 3.35, with 3.01 for the confidence limit closest to the null, quantifying the minimum strength of association that an unmeasured confounder would need to have with both exposure and mortality, on the risk-ratio scale and under the E-value assumptions, to fully explain the observed association (VanderWeele & Ding 2017). Together with the extensive covariate balance achieved in the DAG-informed analyses, these findings indicate that the measured demographic, behavioral, socioeconomic, geographic, pigmentation-related, and baseline health factors do not account for the severe-deficiency mortality association.

### 3.8 Raw and nadir-normalized 25(OH)D provide distinct prognostic information

We compared otherwise identical Cox models containing clinical covariates only, covariates plus raw vitamin D category, or covariates plus nadir-equivalent vitamin D category. Nadir-based models had lower AIC than raw-vitamin-D models for 17 of 19 pooled outcomes; for mortality, AIC improved by 103.8 points (raw minus nadir). Figure 5 shows time-dependent AUC trajectories for nine representative outcomes: overall mortality, diabetes mellitus, diabetic retinopathy, schizophrenia, myocardial infarction, congestive heart failure, cirrhosis, melanoma, and other skin malignancy. Across the representative outcomes, nadir-normalized 25(OH)D provided discrimination similar to or slightly greater than raw 25(OH)D and generally yielded substantially better model fit than the raw measurement. Both raw and seasonally normalized 25(OH)D improved model fit and discrimination compared with models containing the clinical covariates alone.

**Figure 5.**
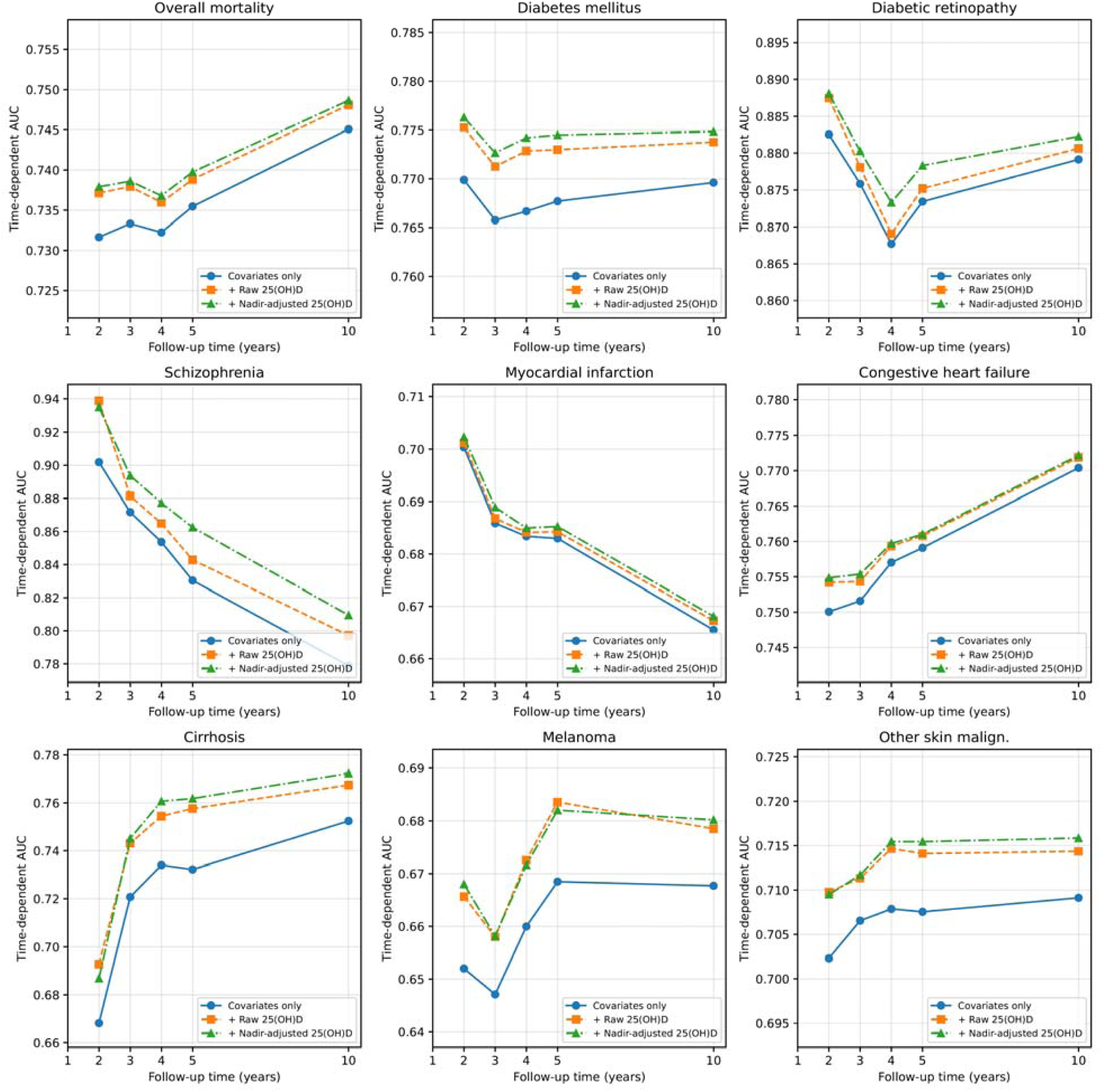
Incremental discrimination contributed by raw and nadir-adjusted 25(OH)D. Time-dependent area under the receiver operating characteristic curve (AUC) is shown at 2, 3, 4, 5, and 10 years for models containing clinical covariates alone, covariates plus raw 25(OH)D category, and covariates plus nadir-adjusted 25(OH)D category. The nine representative outcomes shown are overall mortality, diabetes mellitus, diabetic retinopathy, schizophrenia, myocardial infarction, congestive heart failure, cirrhosis, melanoma, and other skin malignancy. Across the representative outcomes, nadir-normalized 25(OH)D provided discrimination similar to or slightly greater than raw 25(OH)D and generally yielded substantially better model fit than the raw measurement. Both raw and seasonally normalized 25(OH)D improved model fit and discrimination compared with models containing the clinical covariates alone.

## 4. Discussion

### 4.1 Principal findings

In this prospective analysis of over 400,000 UK Biobank participants, seasonally normalized vitamin D status reproduced and extended the systemic pattern observed previously in LHS and TriNetX (Israel, Weizman, et al. 2026). Very severe deficiency below 5 ng/mL was associated with approximately twice the adjusted mortality hazard, while associations were independently reproduced for diabetes, diabetic retinopathy, myocardial infarction, cerebrovascular disease, dementia, and end-stage renal disease. The strongest mortality excess occurred below 10 ng/mL, particularly below 5 ng/mL, whereas little additional mortality reduction was evident above approximately 15–20 ng/mL when vitamin D was expressed at the estimated annual nadir.

The present study adds three distinct layers to the preceding work. First, it validates the findings in a third population with different recruitment and ascertainment. Second, it uses deep phenotyping to characterize and balance major determinants of vitamin D status that were incompletely available in healthcare databases. Third, it treats calendar season as an exposure-measurement problem rather than merely another covariate, demonstrating that almost one half of the cohort changes category when an isolated measurement is translated to a common seasonal reference. A fourth layer is provided by the discrimination analyses: vitamin D adds information beyond the measured covariates across representative systemic outcomes.

### 4.2 Convergence with LHS and TriNetX strengthens the argument for modifiability

In our previous study conducted in parallel within Leumit Health Services (LHS) in Israel and the US TriNetX federated network, severe vitamin D deficiency was associated with mortality and concordant multisystem outcomes in two independent healthcare populations. In LHS, we additionally reconstructed vitamin D supplementation longitudinally from pharmacy dispensing data. Dispensed purchases were calibrated against subsequent measured 25(OH)D responses and modeled as a time-dependent exposure. Supplementation remained independently and dose-dependently associated with lower mortality after adjustment for baseline vitamin D status, and the association remained directionally consistent in a marginal structural sensitivity analysis.

The UK Biobank analysis approaches the same causal question from a complementary direction. Although it does not provide the longitudinal supplementation-purchase data available in LHS, it offers unusually detailed characterization of pigmentation, outdoor exposure, geography, ethnicity, deprivation, smoking, adiposity, and baseline health. When these measured factors were used to construct a covariate-overlap population, very severe deficiency retained an approximately twofold mortality association. The convergence of cross-population replication, dose-response, longitudinal supplementation findings, landmark analyses, and measured-confounder balancing substantially strengthens the evidence for a biological contribution of vitamin D status.

### 4.3 Convergence with randomized and genetic evidence

The present findings should be considered alongside randomized and genetic evidence. Large population-based supplementation trials, including VITAL and D-Health, did not demonstrate reductions in all-cause mortality, but these trials enrolled largely unselected populations rather than participants selected for severe vitamin D deficiency. This distinction may be important given that the strongest associations in our analyses were concentrated below 10 ng/mL, particularly below 5 ng/mL. A recent UK Biobank trial-emulation analysis reproduced the null results of major supplementation trials under comparable vitamin D distributions, while estimating greater benefit when analyses were restricted to participants with vitamin D insufficiency or deficiency. Complementarily, nonlinear Mendelian-randomization analyses in UK Biobank demonstrated an L-shaped association between genetically predicted 25(OH)D and mortality, with risk decreasing steeply at low concentrations and little additional reduction above approximately 50 nmol/L (20 ng/mL). This threshold closely parallels the range in which the strongest mortality gradient was observed in the present study (Manson et al. 2019; Neale et al. 2022; Sutherland et al. 2022; Wang et al. 2026).

### 4.4 Calendar adjustment of vitamin D measurements

Seasonal variation of 25(OH)D has long been recognized, notably in British populations (Hyppönen & Power 2007), and we observed the same pattern in Israel. The present analysis shows why this matters for threshold-based epidemiology. In the fitted UK Biobank trajectory, an observed 25(OH)D concentration of 20.0 ng/mL near the summer peak corresponds to a nadir-equivalent value of approximately 12.1 ng/mL, whereas the same observed value near the late-winter nadir corresponds to approximately 20.0 ng/mL and would be expected to rise to about 33.3 ng/mL in summer. Thus, identical measured concentrations can represent markedly different year-round vitamin D states.

If adverse outcomes are driven particularly by periods of severe deficiency, the lower portion of the annual 25(OH)D trajectory may be more etiologically relevant than an arbitrarily timed measurement. Seasonal normalization therefore addresses exposure misclassification before category assignment, rather than merely adjusting for month or season in the outcome model. This distinction was substantial: 49.2% of participants changed vitamin D category after normalization, rising from approximately 4% near the nadir to >80% around the seasonal peak.

These findings may help explain variation in reported vitamin D thresholds across observational studies and alter interpretation of the plateau observed here. The apparent stabilization of mortality above approximately 15–20 ng/mL was observed here for nadir-equivalent 25(OH)D, rather than for a measurement obtained at an arbitrary time of year.

An interactive implementation of this normalization is available at https://vitamind.medil.ai.

### 4.5 Conventional regression and causal weighting answer complementary questions

The conventional multivariable Cox models and generalized overlap-weighted analysis estimate different quantities in different target populations. Conventional models characterize the full dose-response, sex-specific heterogeneity, and associations across multiple disease domains, while the overlap-weighted analysis tests whether mortality differences persist among participants with substantial covariate overlap after balancing measured common causes of exposure and outcome (Li et al. 2018; Li & Li 2019). Overlap weighting was particularly appropriate because conventional inverse-probability weights can become unstable when exposure probabilities approach zero (Cole & Hernán 2008).

The marked post-weighting reduction in SMDs showed that major baseline differences in ethnicity, pigmentation, deprivation, outdoor exposure, age, smoking, and other factors were effectively balanced, yet the severe-deficiency mortality association persisted. These DAG-informed analyses therefore support a causal interpretation. Together with the independent longitudinal supplementation findings from LHS, stability across progressively longer landmark analyses, and the E-value sensitivity analysis, they substantially strengthen the evidence that severe vitamin D deficiency contributes biologically to increased mortality risk.

### 4.6 The ultraviolet–vitamin D trade-off

One informative finding of both the present and our previous study is that skin malignancies move in the opposite direction from mortality and systemic disease. Ultraviolet radiation is simultaneously the major environmental driver of cutaneous vitamin D synthesis and a cause of cumulative photodamage and skin cancer (Armstrong & Kricker 2001; Gandini et al. 2005). Accordingly, higher vitamin D status can carry the environmental signature of greater ultraviolet exposure even when vitamin D itself is not the cause of the skin-cancer association.

This counter-gradient functions both as a negative control against a nonspecific tendency of low 25(OH)D to predict adverse outcomes generally and as a positive photobiological control for ultraviolet-related exposure. Its persistence after adjustment for measured UV-related covariates indicates that circulating 25(OH)D retains information about ultraviolet exposure not fully captured by questionnaire and geographic measures, while its opposite direction from systemic outcomes argues against a simple nonspecific artifact.

### 4.7 Evolutionary implications and a testable host-photobiology framework

Human skin pigmentation shows strong geographic adaptation to ultraviolet radiation, which can be interpreted as an evolutionary balance between photoprotection under high UV exposure and sufficient UVB penetration for cutaneous vitamin D synthesis in lower-UV environments (Chaplin & Jablonski 2009; Jablonski & Chaplin 2010). Our present findings extend this framework to overall mortality and long-term health outcomes. If prolonged vitamin D deficiency compromises survival and multisystem health, while elevated UV exposure accelerates cumulative photodamage, natural selection should favor phenotypes that optimize vitamin D synthesis while minimizing the complications of excess UV exposure. The opposing gradients observed in this study—elevated mortality and systemic disease risk associated with low vitamin D status, contrasted with a lower risk of skin malignancies—provide strong empirical evidence of this biological trade-off.

Skin pigmentation is the most obvious component of this photobiological system, but it may not be the only one. Hair characteristics may also modify the fraction of ambient ultraviolet radiation reaching the scalp; experimental studies show that hair attenuates both UVA and UVB, with protection varying by density, thickness, and pigmentation. Decomposing this host-photobiology node represents a mechanistic question for future investigation.

### 4.8 Potential immune and infectious mechanisms

The biological mechanisms linking severe vitamin D deficiency to such a broad range of outcomes remain incompletely understood. The replicated associations across metabolic, vascular, neurological, microvascular, renal, and mortality outcomes are compatible with mechanisms acting at the level of systemic host resilience rather than through multiple unrelated disease-specific pathways. Potential mechanisms include chronic inflammation and immune dysregulation (Hewison 2011; Schwalfenberg 2011), endothelial dysfunction (Kim et al. 2020), and altered host–microbial interactions or impaired containment of persistent infection (Lagishetty et al. 2011; Ismailova & White 2022). Within this framework, we recently proposed that persistent *Toxoplasma gondii* infection, an extremely prevalent chronic protozoal infection, may contribute to late-life morbidity through delayed pathogenicity, and that vitamin D deficiency could facilitate this process by creating a less effective immune environment for containment of infection (Israel & Israel 2026; Israel, Merzon, et al. 2026). The present UK Biobank analysis does not directly assess *T. gondii* infection and therefore cannot test this mechanism. Nevertheless, the reproducible association of severe vitamin D deficiency with outcomes across several organ systems is compatible with a shared immune or host–pathogen pathway and provides a rationale for future studies integrating vitamin D status with serological or molecular markers of persistent infection and immune activation.

### 4.9 Strengths and limitations

This study has several major strengths. First, it examines vitamin D status in one of the largest deeply phenotyped prospective cohorts available, with more than 400,000 participants contributing to the principal analyses, long longitudinal follow-up, and more than 31,000 deaths. Second, UK Biobank provides systematic baseline characterization of major determinants of vitamin D status and mortality, including pigmentation, ethnicity, outdoor exposure, geography, socioeconomic deprivation, smoking, adiposity, fish-oil uptake, and baseline morbidity. Use of complete-case analyses ensured that all prespecified measured confounders were directly observed for included participants and allowed extensive adjustment without model-based imputation. This enabled substantially more extensive control of measured confounding than was possible in electronic-health-record cohorts.

Third, the study independently reproduced the mortality and multisystem disease associations previously observed in two very different healthcare populations in Israel and the United States. This cross-population consistency is particularly notable because UK Biobank participants differ markedly from the LHS and TriNetX populations in recruitment and ascertainment, and are, on average, healthier and less socioeconomically deprived than the general UK population. The persistence of strong associations in this relatively healthy volunteer cohort therefore strengthens the robustness and external reproducibility of the findings.

Fourth, the study explicitly addressed seasonal exposure. The direct comparison of raw and seasonally normalized exposure definitions showed consistently improved model fit, although gains in discrimination were modest, supporting improved exposure specification rather than a large increase in predictive performance.

Fifth, causal inference was approached using a prespecified DAG, generalized overlap weighting, detailed balance diagnostics, progressively longer landmark exclusions, E-value sensitivity analysis, and geographic sensitivity analyses. Overlap weighting achieved strong balance across an extensive measured covariate set while the severe-deficiency mortality association remained approximately twofold. The stability of estimates across landmark exclusions extending to 5 years further argues against early occult disease or short-term reverse causation as the principal explanation.

Sixth, melanoma and other skin malignancies provided informative control outcomes. As in the preceding LHS and TriNetX analyses, these outcomes moved in the opposite direction from mortality and systemic disease, consistent with ultraviolet-related biology. This counter-gradient argues against a nonspecific tendency for low 25(OH)D to associate with every adverse outcome and provides an internal positive control that the exposure retains coherent information about ultraviolet exposure.

Together, these complementary features provide strong observational support for a biological contribution of severe vitamin D deficiency to mortality and multisystem vulnerability. Several limitations remain. First, vitamin D status was derived from a single baseline measurement. Seasonal normalization adjusts for the predictable population-level annual component but cannot reconstruct each participant’s true longitudinal vitamin D trajectory, and individuals may differ in the amplitude and shape of seasonal variation. Second, supplementation information in UK Biobank was substantially less detailed than in LHS, and available baseline supplement variables were comparatively coarse.

Despite the extensive phenotyping available in UK Biobank, residual unmeasured confounding cannot be entirely excluded. However, the broad measured covariate set, strong balance achieved after overlap weighting, E-value analysis, and stability across progressively longer delayed-entry lags substantially narrow the range of plausible confounding explanations. Some secondary outcomes were uncommon, reducing precision and occasionally leading to numerically unstable proportional-hazards diagnostics in sparse sex-specific analyses; the corresponding outcome estimates remained estimable, and exposure-focused diagnostics were examined separately.

## 5. Conclusions

In over 400,000 deeply phenotyped UK Biobank participants, seasonally normalized severe vitamin D deficiency was strongly associated with mortality and with major metabolic, vascular, neurological, microvascular, and renal outcomes previously identified in independent Israeli and US populations. The strongest mortality excess occurred below 10 ng/mL nadir-equivalent 25(OH)D, particularly below 5 ng/mL, and persisted after generalized overlap weighting and progressively longer landmark exclusions. Together with the preceding longitudinal supplementation findings, these results support severe vitamin D deficiency as a potentially modifiable contributor to aging-related systemic vulnerability.

Seasonal normalization may provide a more biologically meaningful framework for future vitamin D epidemiology, while the opposing gradients of systemic disease and ultraviolet-related skin malignancy highlight the broader trade-off between effective ultraviolet exposure, vitamin D physiology, and photoprotection. These population-scale findings provide a framework for future mechanistic and interventional studies, including investigation of immune and host–microbial pathways.

## Data Availability

The data supporting the findings of this study are available to approved researchers through UK Biobank following its standard application and access procedures. The present study was conducted under UK Biobank Application Number 56774. Participant-level data cannot be redistributed by the authors.

https://www.ukbiobank.ac.uk/

https://vitamind.medil.ai/

## Acknowledgements

This research has been conducted using the UK Biobank Resource under Application Number 56774.

## Author contributions

Conceptualization: AI, EMe, NR Methodology

AI, NR Investigation: AI, NR, EMe Writing – original draft: AI, NR

Writing – review & editing: AI, AW, SA, SV, EMa, EMe

## Materials & Correspondence

Correspondence and requests for materials should be addressed to the corresponding author.

## Abbreviations

25(OH)D: 25-hydroxyvitamin D
AIC: Akaike information criterion
AUC: Area under the receiver operating characteristic curve
BMI: Body mass index
CI: Confidence interval
DAG: Directed acyclic graph
HR: Hazard ratio
ICD-10: International Classification of Diseases, 10th Revision
LHS: Leumit Health Services
SMD: Standardized mean difference
UV: Ultraviolet
UVA: Ultraviolet A
UVB: Ultraviolet B

