## Supplementary Material for "Seasonally Normalized Vitamin D Status and Long-Term Mortality and Multisystem Disease Risk in the UK Biobank"

### Supplementary Methods

#### Cohort assembly and baseline data extraction

UK Biobank baseline data were assembled from the first assessment instance (station 0 where applicable). The extracted baseline cohort initially contained 501,936 participants. The extraction retained participants with non-missing age (field 21003) and a baseline assessment year of 2007 or later (n=498,147); the source cohort comprised the 447,842 participants with an available baseline serum 25-hydroxyvitamin D [25(OH)D] measurement (field 30890). Baseline assessment date was obtained from field 53 and assessment centre from field 54.

#### Vitamin D measurement and seasonal normalization

Baseline serum 25(OH)D was obtained from UK Biobank field 30890 and converted from nmol/L to ng/mL by division by 2.5. Calendar day of measurement was represented by eight sine/cosine harmonic pairs. With ω = 2π/365.25, the fitted seasonal component was S(d) = Σ(k=1,…,8){βs,k sin(kωd) + βc,k cos(kωd)}. Coefficients were estimated by linear regression of log-transformed baseline 25(OH)D on the 16 harmonic terms in the full vitamin D cohort. The fitted annual nadir occurred on day 59 (late February). For participant i measured on day d_i, the nadir-equivalent concentration was calculated as exp{log[25(OH)D_observed,i] − S(d_i) + S(d_nadir)}. This removes the fitted calendar component from the observed measurement and expresses each value at the common late-winter reference.

The submitted analyses used the following harmonic coefficients: sin1 −0.19640784, cos1 −0.17189752; sin2 0.01193693, cos2 0.00768729; sin3 0.01205368, cos3 −0.00959092; sin4 −0.00579821, cos4 0.00407145; sin5 −0.00099733, cos5 −0.00657994; sin6 −0.00196533, cos6 −0.00320386; sin7 −0.00183577, cos7 −0.00400819; and sin8 −0.00385140, cos8 −0.00265219. These coefficients correspond to the normalization model used to generate the submitted nadir-equivalent exposure.

#### Baseline covariates

The main association models used age, sex, broad self-reported ethnicity, BMI category and within-category BMI offset, baseline diabetes, current smoking, past smoking, skin colour, reported summer and winter outdoor exposure, Townsend deprivation index, geographic position, calendar year of assessment, and fish-oil use. Sex was omitted from sex-specific models. The UK Biobank source fields and preprocessing rules used in the extraction are summarized below.

| Variable | UK Biobank field | Extraction / recoding used |
| --- | --- | --- |
| Baseline 25(OH)D | 30890 | Converted from nmol/L to ng/mL by division by 2.5. |
| Baseline assessment date | 53 | Parsed as date; used to derive calendar year, month and day of year. |
| Assessment centre | 54 | Retained for geographic sensitivity analyses. |
| Age | 21003 | Numeric; non-missing age required in cohort preprocessing. |
| Sex | 31 | Mapped 0 = female, 1 = male. |
| BMI | 21001 | Numeric; subsequently categorized with a within-category BMI offset. |
| Self-reported ethnic background | 21000 | Mapped using UKB coding and collapsed to broad ethnicity groups for modeling. |
| Current smoking | 1239 | 0 = no smoker, 1 = smoker, 2 = occasional smoker; missing mapped to no smoker in preprocessing. |
| Past smoking | 1249 | Code 1 = past smoker; code 4 = never smoker; other/missing responses retained as a residual category. |
| Skin colour | 1717 | Mapped using UKB coding; missing assigned to “Do not know” in preprocessing. |
| Summer outdoor time | 1050 | Codes −1/−3 treated as missing; −10 represented as 0.5 h/day. |
| Winter outdoor time | 1060 | Codes −1/−3 treated as missing; −10 represented as 0.5 h/day. |
| Townsend deprivation | 22189 | Numeric value used as Townsend deprivation index in the extraction. |
| geographic latitude | 54 | Z-standardized latitude of the primary assessment centre. |
| Fish-oil supplement use | 6179 | Baseline multi-select field; code 1 indicates fish-oil use. |
| Medication codes | 20003 | Searched for vitamin D/calciferol medication codes; no positive observations in extraction. |
| Baseline diabetes dates | 130706 / 130708 | E10/E11 first-report dates; baseline diabetes if date ≤ baseline assessment. |

BMI was grouped using cut points 10, 18.5, 25, 30, 35, 40, 45 and 99 kg/m², with 18.5–25 kg/m² as the reference category. A within-category BMI offset was calculated as the difference between measured BMI and the lower boundary of its category. Baseline diabetes was defined by an E10 or E11 first-report date (fields 130706 and 130708) on or before the baseline assessment.

Self-reported ethnic background (field 21000) was mapped using UK Biobank coding and subsequently collapsed for the causal analyses into White, Asian, Black, Mixed, Chinese and Other; responses indicating “prefer not to answer,” “do not know,” or unknown ethnicity were treated as missing. Current smoking (field 1239) was represented as no smoking, smoking, or occasional smoking. Past smoking (field 1249) identified code 1 as past smoker and code 4 as never smoker; other or missing responses were retained as a residual category. Missing smoking information was uncommon, affecting fewer than 0.2% of participants; during preprocessing, these values were assigned to the non-current-smoker reference category.

Skin colour (field 1717) was mapped using the corresponding UK Biobank coding, with missing values assigned to “Do not know” in preprocessing. For summer and winter outdoor time (fields 1050 and 1060), special codes −1 and −3 were treated as missing and −10 (“less than an hour a day”) was represented as 0.5 h/day. Fish-oil use was derived from baseline multi-select supplement field 6179 (code 1). Vitamin D-containing medications were additionally sought in baseline medication field 20003 by matching medication-dictionary entries containing “vitamin D” or “calciferol”; no positive observations were identified during preprocessing, and this variable therefore did not enter the fitted models.

Geographic position was represented by the z-standardized latitude of the assessment centre, derived from UK Biobank field 54.. Calendar year and day of year were derived from the baseline assessment date (field 53), with calendar year represented in modeling as years since 2006. Townsend deprivation was extracted from field 22189, as used in the preprocessing dataset.

#### Outcome ascertainment

For non-cancer outcomes, the earliest available date for each selected UK Biobank first-report field was extracted from the date dataset. For cancer outcomes, the earliest cancer-registry diagnosis date was identified after truncating ICD-10 codes to three characters. Composite outcomes used the earliest component date. An outcome was considered incident when its first recorded date occurred after the baseline assessment; the primary analyses then applied the prespecified one-year delayed-entry restriction. Full operational definitions and source fields are provided in Supplementary Table S1.

#### Follow-up, censoring and delayed entry

Time zero was the baseline UK Biobank assessment/25(OH)D measurement (field 53). For each outcome, follow-up ended at the earliest of the outcome date, death for nonfatal outcomes (field 40000), recorded loss to follow-up (field 191), or the global data-censoring date, defined in the analysis as one day after the latest recorded death date in the dataset. Follow-up was additionally capped at 13.1 years from each participant’s baseline assessment. The primary analyses used delayed entry at one year after baseline, retaining baseline as the time origin; participants with an event or censoring before delayed entry did not enter that analysis. Reverse-causation sensitivity analyses repeated the full causal procedure using delayed-entry lags of 0, 1, 2, 3, 4 and 5 years.

#### Conventional association models and complete-case preparation

Cox proportional-hazards models were fitted for all-cause mortality and cause-specific Cox models for each nonfatal outcome, with death treated as a censoring event for nonfatal outcomes. Model-specific complete-case datasets were constructed from the exposure, outcome/time variables and prespecified covariates. Covariates with fewer than two observed levels after complete-case restriction were removed iteratively, after which the complete-case population was rebuilt so that missingness in a discarded covariate did not unnecessarily exclude participants. Adjusted hazard ratios and 95% confidence intervals were estimated for each nadir-equivalent 25(OH)D category relative to the ≥20 ng/mL reference category.

Proportional-hazards assumptions were assessed first using scaled Schoenfeld residuals from the full adjusted model. If the full diagnostic was numerically singular, a full-model vitamin D-by-log(time) interaction test was used; only if that also failed was a reduced nuisance-covariate Schoenfeld diagnostic used to assess time variation of the vitamin D effect.

#### Raw-versus-nadir model comparison and discrimination

For each outcome and sex stratum, covariate-only, raw-vitamin-D and nadir-equivalent-vitamin-D Cox models were fitted on the identical complete-case population. Model fit was compared using Akaike information criterion and concordance. Time-dependent AUC accounting for right censoring was calculated from the baseline time origin at 2, 3, 4, 5 and 10 years. The 1-year horizon was not reported because eligibility for the primary analysis required remaining event-free and under follow-up through the one-year delayed-entry point. Overall and month-specific movement between raw and nadir-equivalent categories was also quantified.

#### DAG-informed causal mortality analysis

The causal mortality adjustment set comprised the main association covariates plus baseline histories of myocardial infarction, congestive heart failure, cerebral infarction, chronic kidney disease/end-stage renal disease, cirrhosis, dementia and cancer. Baseline disease histories were defined by a relevant first-report or cancer-registry date on or before the baseline assessment. Incident outcomes potentially downstream of vitamin D status were not entered as time-varying mortality covariates.

Generalized propensity scores for the five vitamin D categories were estimated by multinomial regression. Generalized overlap weights were calculated as h(x)/e_k(x), with h(x)=1/sum_j[1/e_j(x)], and were rescaled to a mean of 1. Weighted Cox models used robust variance estimation clustered by participant identifier. Covariate balance was assessed using the maximum absolute pairwise standardized mean difference for each covariate across treatment groups, with <0.10 prespecified as acceptable. Propensity-score distributions, weight distributions and effective sample sizes were recorded as diagnostics.

Weighted survival functions were used to derive absolute mortality risks at 5 and 10 years from baseline and corresponding risk differences and risk ratios across vitamin D categories. Additional causal sensitivity analyses compared <10 ng/mL with >20 ng/mL and >30 ng/mL, replaced the primary geographic variable with assessment centre, and, where cell sizes permitted, replaced broad ethnicity with the original self-reported ethnicity categories.

A calibration-refined stabilized multinomial IPTW analysis was implemented as a sensitivity analysis. Stabilized IPTW weights were trimmed at the 1st and 99th percentiles and then calibrated within vitamin D groups to the pooled covariate means. This sensitivity estimate was prespecified for manuscript use only if the maximum post-calibration pairwise SMD was <0.10.

E-values were calculated from the overlap-weighted hazard ratio and the confidence limit closest to the null. For HRs above 1, E = HR + sqrt[HR×(HR−1)]; for protective estimates, the reciprocal HR was used. These values quantify the minimum association strength an unmeasured confounder would require with both exposure and outcome, on the risk-ratio scale under the E-value assumptions, to explain the observed association.

#### Ultraviolet-related control-outcome analyses

Melanoma and other skin malignancies were examined as photobiological control outcomes. Nested models compared a core adjustment set with a set additionally including skin colour, summer and winter outdoor exposure and geographic position, to assess whether the inverse skin-cancer gradient persisted after measured ultraviolet-related covariates were included.

#### Multiplicity and descriptive analyses

Continuous variables were summarized as mean and standard deviation where appropriate and categorical variables as counts and percentages. Standardized mean differences were emphasized for baseline comparisons because of the large cohort size. For the secondary outcome analyses, Benjamini–Hochberg false-discovery-rate adjustment was applied across non-reference vitamin D contrasts. An ordered-category trend test used the five vitamin D categories scored from lowest to highest.

### Supplementary Table S1. Operational definitions of study outcomes

Non-cancer first-report dates were taken from the UK Biobank date dataset; cancer outcomes used the earliest cancer-registry diagnosis date for the indicated three-character ICD-10 code(s). Composite outcomes used the earliest component date. The primary analyses applied one-year delayed entry after baseline.

| **Outcome** | **Operational definition** | **UK Biobank source / field(s)** | **Notes** |
| --- | --- | --- | --- |
| All-cause mortality | Recorded death date | UKB field 40000 (death_date) | Primary outcome. |
| Diabetes mellitus | Earliest E10 or E11 first-report date | Fields 130706 (E10), 130708 (E11) | Notebook notes that E10/E11 distinction was considered unreliable for the composite DM outcome. |
| Type 2 diabetes | E11 first-report date | Field 130708 (E11) | Secondary analysis; participants with baseline diabetes were excluded in the R script. |
| Diabetic retinopathy | H36 first-report date | Field 131184 (H36) | Implemented in the analysis as diabetic retinopathy. |
| Myocardial infarction | I21 first-report date | Field 131298 (I21) |  |
| Congestive heart failure | I50 first-report date | Field 131354 (I50) |  |
| Cerebral infarction / ischemic stroke | I63 first-report date | Field 131366 (I63) | Analysis variable is labelled CVA in the R script; I63 is the implemented definition. |
| Hemiplegia | G81 first-report date | Field 131102 (G81) |  |
| Schizophrenia | F20 first-report date | Field 130874 (F20) |  |
| Cirrhosis | K74 first-report date | Field 131666 (K74) |  |
| Dementia | Earliest F00 or F03 first-report date | Fields 130836 (F00), 130842 (F03) | G30 was available for baseline-history construction but was not part of the incident dementia outcome definition. |
| Chronic kidney disease | N18 first-report date | Field 132032 (N18) |  |
| End-stage renal disease | Date of end-stage renal disease report | Field 42026 (ESRD) |  |
| Hearing loss | H90 first-report date | Field 131258 (H90) |  |
| Gastrointestinal malignancy | Earliest cancer-registry diagnosis with ICD-10 C15–C26 | Cancer registry (date_diagnosis); C15–C26 | Three-character ICD-10 codes. |
| Lung malignancy | Earliest cancer-registry diagnosis with ICD-10 C34 | Cancer registry (date_diagnosis); C34 |  |
| Colon cancer | Earliest cancer-registry diagnosis with ICD-10 C18 | Cancer registry (date_diagnosis); C18 | Analysis variable is labelled CRC in the R script, but the implemented code is C18 only. |
| Melanoma | Earliest cancer-registry diagnosis with ICD-10 C43 or D03 | Cancer registry (date_diagnosis); C43 or D03 | Includes invasive melanoma and melanoma in situ. |
| Other skin malignancy | Earliest cancer-registry diagnosis with ICD-10 C44 | Cancer registry (date_diagnosis); C44 | Used as an ultraviolet-related photobiological control outcome. |
